# Arm-hand boost therapy during inpatient stroke rehabilitation: a pilot randomized controlled trial

**DOI:** 10.1101/2021.01.12.21249586

**Authors:** Sarah Meyer, Geert Verheyden, Kristof Kempeneers, Marc Michielsen

## Abstract

**Objective:** It was the aim to assess feasibility, safety and potential efficacy of a new intensive, focused arm-hand BOOST program and to investigate whether there is a difference between early versus late delivery of the program in the sub-acute phase post stroke.

**Methods:** In this pilot RCT, patients with stroke were randomized to the immediate group (IG): 4 weeks (4w) BOOST +4w CONTROL or the delayed group (DG): 4w CONTROL +4w BOOST, on top of their usual inpatient care program. The focused arm-hand BOOST program (1 hour/day, 5x/week, 4 weeks) consisted of group exercises with focus on scapula-setting, core-stability, manipulation and complex ADL tasks. Additionally, one hour per week the Armeo^®^Power (Hocoma AG, Switzerland) was used. The CONTROL intervention comprised a dose-matched program (24 one-hour sessions in 4w) of lower limb strengthening exercises and general reconditioning. At baseline, after 4 and 8 weeks of training, the Fugl-Meyer assessment upper extremity (FMA-UE), action research arm test (ARAT) and stroke upper limb capacity scale (SULCS) were administered.

**Results:** Eighteen participants (IG: n=10, DG: n=8) were included, with a median (IQR) time post stroke of 8.6 weeks (5-12). No adverse events were experienced. After 4 weeks of training, significant between-group differences were found for FMA-UE (p=0.003) and SULCS (p=0.033) and a trend for ARAT (p=0.075) with median (IQR) change scores for the IG of 9 (7-16), 2 (1-3) and 12.5 (1-18) respectively, and for the DG of 0.5 (−3-3), 1 (0-1) and 1.5 (−1-9), respectively. In the IG, 80% of patients improved beyond the minimal clinical important difference of FMA-UE after 4 weeks, compared to none of the DG patients. Between 4 and 8 weeks of training, patients in the DG tend to show larger improvements when compared to the IG, however, between-group comparisons did not reach significance.

**Conclusions:** Results of this pilot RCT showed that an intensive, specific arm-hand BOOST program, on top of usual care, is feasible and safe in the sub-acute phase post stroke and suggests positive, clinical meaningful effects on upper limb function, especially when delivered in the early sub-acute phase post stroke.

**Clinical Trial Registration:** www.ClinicalTrials.gov, identifier NCT04584177

## 1 Introduction

Approximately 70% of stroke survivors experience impairments in the upper limb (1). As a consequence, upper extremity functions, such as reaching, grasping, releasing and manipulating objects are hindered, often resulting in a learned non-use of the affected upper limb (2). Dysfunction in the upper limb post stroke can therefore significantly limit a person’s level of activity and quality of life (3). The importance of upper limb treatment is acknowledged by patients with stroke, their caregivers and different health professionals who agreed upon the top 10 research priorities relating to life after stroke (4,5). An early start of arm and hand treatment is crucial when considering the critical window of opportunity for recovery when the brain is most responsive to sensorimotor experience after injury, typically seen in the first 3 months post stroke (6-9).

In 2015, the rehabilitation team in Jessa Hospital, Belgium developed the JSU (Jessa Sint Ursula)-diagram, with the objective to stimulate the early start of rehabilitation of the arm and hand post stroke (10). The diagram offers a guideline to work on the objectives needed to shape the rehabilitation of the upper limb at various stages of recovery. The diagram also highlights the importance of stratification of patients towards different training objectives, based on the levels of trunk control and arm function that are reached (10). A focused therapy program for the upper limb needs to be further delineated by knowledge of neurophysiological recovery post stroke to ensure that specific therapy goals are set for the correct patient at each stage in recovery (11-13). The prerequisites for recovery of voluntary selective movements of the upper limb in patients post stroke include a large proximal component such as adequate postural control and core stability (14,15), correct scapula setting (16,17), efficient scapular humeral rhythm (17-19) and selective recruitment of reach-related musculature (20,21). The importance of the proximal component in arm hand rehabilitation was already shown by Feys et al. who found a clinically meaningful and long-lasting effect up to five years post stroke of an early, repetitive stimulation of the shoulder complex by using a rocking chair (22). Besides the proximal component, specific attention is needed towards the rehabilitation of the hand, including restoring the different arches of the hand (23) to provide a stable base and correct alignment as a prerequisite for dexterity (24) and hand shape modulation when reaching to objects (25).

In addition to the importance of focused upper limb therapy that can maximize neurophysiological recovery post stroke, considerable agreement (26,27) exists about the importance of high dose (time in rehabilitation, or amount of repetitions) and intensity (dose per session) of upper limb therapy. In regular therapy, it has been reported that patients make a total of only about 30 upper limb task-based repetitions during a single therapy session, which is not sufficient to expect large improvements (28,29). Very recently, Krakauer et al. (30) reported significant improvements in action research arm test (ARAT) scores, but not in Fugl-Meyer assessment upper extremity (FMA-UE) scores, when intensifying the rehabilitation training program in the sub-acute phase post stroke by using a novel exploratory neuro-animation therapy (30). Other recent studies but in patients with chronic stroke have shown improvements at both the activity and impairment levels when greatly increased intensities and doses of upper limb therapy are provided (31-34).

In summary, the important role of both the content and the intensity of arm and hand therapy for improving upper limb motor function is well established. However, the effect of a more focused program, including stratification, principles of neurophysiological recovery and therapy including proximal control as well as distal alignment, provided at high intensities in the sub-acute phase post stroke remains poorly understood. Therefore, it was the aim of this study to assess the feasibility, safety and potential efficacy of a new intensive arm-hand BOOST program embedded in the inpatient rehabilitation phase post stroke, when compared to a dose-matched program of strengthening exercises for the lower limbs and general reconditioning. Additionally, it was the objective to investigate whether there was a difference in efficacy between early versus late delivery of the BOOST program. We hypothesized that the intensive arm-hand BOOST program is feasible and safe to deliver in the subacute phase post stroke, on top of the usual care program. Additionally, our hypothesis was that the BOOST program is beneficial for improving upper limb motor function and activities due to the combination of high intensity and specificity of the program. It is further expected that early delivery of the program is better when compared to late delivery post stroke, due to the enlarged sensitivity for rehabilitative therapy early after stroke, which declines with time (9).

## 2 Materials and methods

### 2.1 Study design

This study is an assessor-blinded, pilot randomized controlled trial. A specific intensive boost program for the upper limb (BOOST) was compared to a dose-matched program of strengthening exercises for the lower limbs and general reconditioning (CONTROL). Both interventions were provided in addition to conventional patient-focused inpatient rehabilitation (USUAL CARE).

Eligible patients were block-randomized with an allocation ratio of 1:1 to either the (1) Immediate group (IG): 4 weeks of BOOST, followed by 4 weeks of CONTROL intervention; or (2) Delayed group (DG): 4 weeks of CONTROL intervention, followed by 4 weeks of BOOST therapy. An overview of the study flow diagram is provided in figure 1. The study is reported conform the CONSORT (Consolidated Standards of Reporting Trials) statement (Supplementary Table 1). The study was approved by the ethical review committee of Jessa Hospital on April 2^nd^ 2019 (Number 19.27/Reva19.01, Belgian registration number B243201939920, Chairperson: Dr. Magerman) and conducted in accordance to the principles set forth in the declaration of Helsinki. The trial is registered with clinicaltrials.gov (NCT04584177).

**Figure 1.**
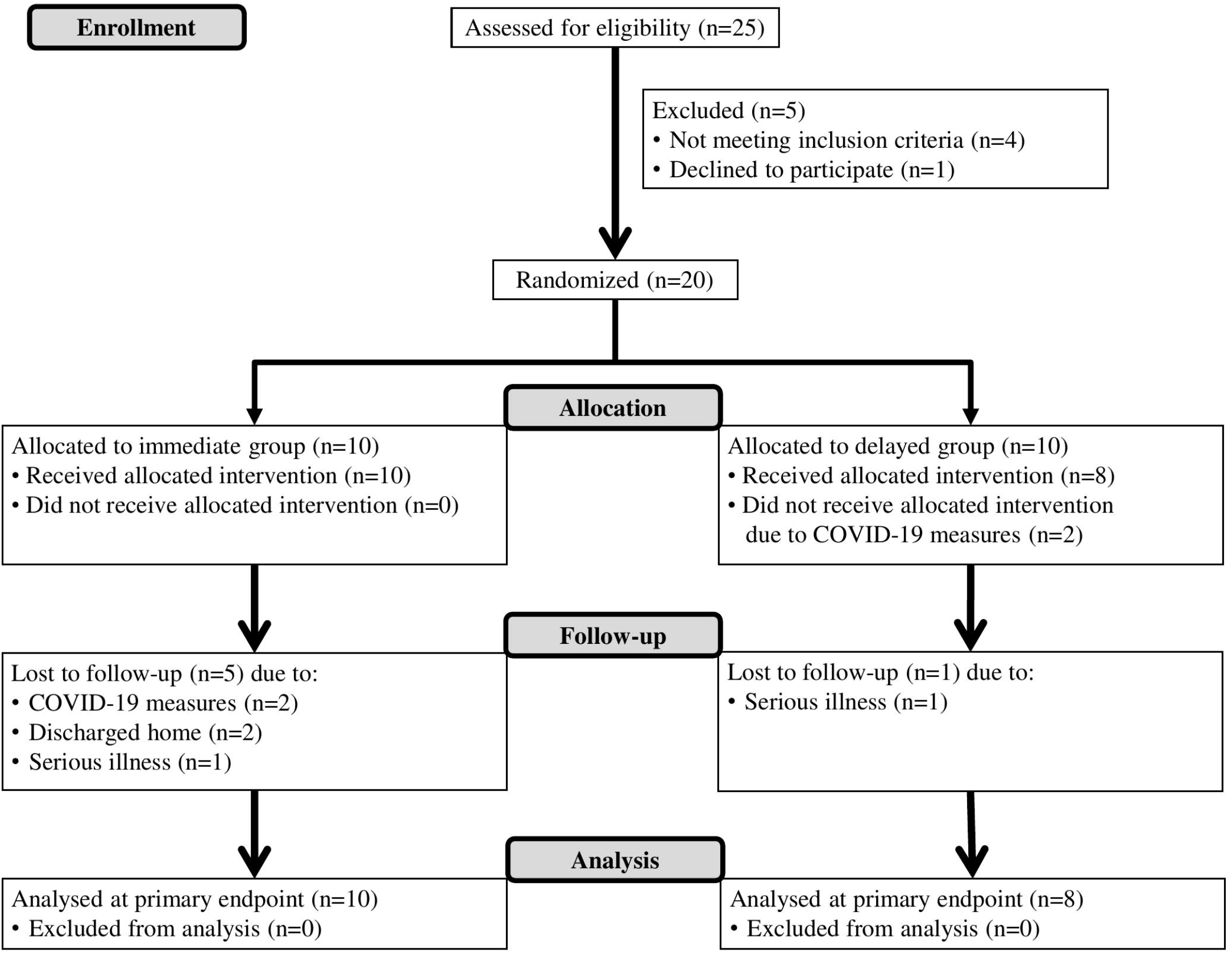
Flow diagram

### 2.2 Participants

Participants were recruited consecutively from the inpatient rehabilitation unit of Jessa Hospital, Rehabilitation Campus Sint-Ursula in Belgium between May 2019 and March 2020. patients were eligible for the study if they (1) experienced a first-ever unilateral, supra-tentorial stroke as defined by the World Health Organization (35), (2) were minimally 18 years old, (3) had a residual inpatient stay of minimally 4 weeks, (4) had the ability to sit independently, as defined as a maximal score of 25 on item 3 of the trunk control test (36), and (5) experienced motor impairment in the upper limb, as defined, based on the JSU diagram (10), as a score of 8-17 on stage 2 (synergies) of the FMA-UE (37), or a score of <8 on stage 2 of the FMA-UE, combined with a score of >6 on stage 5 (hand) of the FMA-UE. The exclusion criteria were: (1) musculoskeletal and/or other neurological conditions with permanent damage that may interfere with the study procedures or assessments, (2) subdural hematoma, tumor, encephalitis or trauma, with stroke-like symptoms, and (3) severe communication or cognitive deficits which could hamper the assessment.

Eligible patients were informed about the content and aims of the study by the team staff members. All patients provided written informed consent prior to participation in the study. An independent researcher (MM) allocated participants to one of the treatment groups, based on a computer-generated list, through informing the treating therapists of the random allocation.

### 2.3 Interventions

The BOOST and CONTROL intervention were provided on top of the conventional patient-focused inpatient rehabilitation. The standard usual care program consists of physiotherapy and occupational therapy, supplemented by speech- and language therapy, psychological therapy and sports therapy. All patients were instructed to refrain from beginning any type of other new treatment for the upper limb during the entire period of the study and to maintain the activities of their normal daily, inpatient lives.

Both the BOOST and CONTROL intervention consist of 20 one-hour group sessions (spread over 4 weeks, 5 days a week) and one hour of individual therapy per week. In total, each of those two study interventions includes 24 hours of additional therapy, provided over a period of 4 weeks. The immediate group (IG) received first 4 weeks of BOOST, followed by 4 weeks of CONTROL intervention; and vice versa for the delayed group (DG): 4 weeks of CONTROL intervention, followed by 4 weeks of BOOST therapy.

#### 2.3.1 BOOST

The specific intensive boost program for the upper limb (BOOST) is focused around 5 topics: scapula-setting, core-stability training in relation to reaching, training of external shoulder rotation and elbow extension (movements with 30-60° flexion/abduction in shoulder), fine manipulation or dexterity training and integration in complex ADL tasks. For each of those topics, a list of example exercises was created that could be used depending on the individual abilities of the patient, including a gradual increase in levels of difficulty. Each of the interventions is tailored to the individual patient, based upon the ongoing assessment using the Model of Bobath Clinical Practice (11), discussion within the group of therapists and individual treatment goals of the patient.

A typical BOOST session would include exercises and tasks with the aim of getting a proper alignment of the upper limb (.. eccentric activation of shortened muscles, realignment of scapulohumeral joint), updating orientation (e.g. light touch, proprioceptive accuracy), activating the upper limb (e.g. scapular stability, weight bearing, activating the intrinsic muscles of the hand, strengthening, reaching, grasping, pinching) and training bilateral limb coordination during functional activities (e.g. cooking/eating, dressing, grooming, cleaning, gardening). The material used in a BOOST session includes standard rehabilitation material such as different sizes of balls, cones, hoops, beads, and daily objects.

The group sessions were executed with a maximum of four participants, under supervision of two experienced therapists (physiotherapist and/or occupational therapist). Additionally, patients exercise one hour per week using the Armeo^®^Power (Hocoma AG, Switzerland), an upper limb exoskeleton device. This allowed practice of 3D arm movements with six actuated degrees-of-freedom. Participants performed exercises at high intensities in the active-assisted mode by using motivational games. A licensed occupational therapist was present throughout each session and provided feedback to encourage typical (non-synergistic) movement patterns at all times.

The intervention is reported conform the TIDieR (Template for intervention description and replication) checklist (Supplementary Table 2).

#### 2.3.2 CONTROL

The CONTROL therapy consists of a program of strengthening exercises for the lower limbs and general reconditioning. During the group sessions, a circuit-class training is performed according to a standardized written protocol: 20 minutes of cycling using the THERA-Trainer tigo^®^ (Thera-Trainer, Germany), 20 minutes of strengthening exercises for muscles around hip and knee (e.g. sit-to-stand training), 10 minutes of knee exercises using the quadriceps bench in free swing mode (Gymna, Belgium) and 10 minutes of leg press exercises using the Minivector^®^ (Easytech, Italy). Again, a gradual increase in levels of difficulty was provided. Group sessions were executed with a maximum of four participants under supervision of one therapist (physiotherapist or occupational therapist). Additionally, each participant received a self-exercise program for the rehabilitation of the lower limbs (balance; strengthening of foot, knee and hip muscles; walking exercises), which was executed one hour per week under minimal supervision of an occupational therapist. None of the exercises involved the use of the upper limb.

### 2.4 Outcome measures

Outcome assessments were performed at three time points: before training (BASELINE), after 4 weeks of training (POST 1) and after 8 weeks of training (POST 2). All assessments were conducted by a trained researcher (SM) who was blinded to treatment allocation. Patients were coached and given a verbal reminder not to speak to the evaluator regarding the therapy type at each assessment visit. The evaluator had no contact with the participants and therapists outside of the assessment sessions to minimize chances of unblinding. At the baseline visit, demographic information was collected such as: age, gender and hand-dominance. Stroke-specific data about the timing, side and type of stroke (ischemic or hemorrhagic) was documented. Patients also underwent a comprehensive clinical evaluation including the Barthel index (38), Montreal cognitive assessment (39) and star cancellation test (40) of visuospatial neglect (presence of hemineglect determined by cutoff score of <44 out of 54).

At the three time points, the following outcome measures were used: Fugl-Meyer assessment upper extremity (FMA-UE) (37), action research arm test (ARAT) (41), stroke upper limb capacity scale (SULCS) (42), box & block test (BBT) (43), Jebsen Taylor hand function test (JEBSEN) (44) and Rivermead motor assessment – arm subscale (RMA-A) (45). The primary outcome measure was the change in upper limb impairment measured by FMA-UE, from BASELINE to POST 1 (after 4 weeks of training). The FMA-UE (37) is a reliable and valid measure of overall motor impairment, widely used in patients with stroke and recommended by the stroke recovery and rehabilitation roundtable for use in stroke recovery trials (46). The scale consists of 33 items graded on an ordinal scale (0-2), with a total score ranging between zero (loss of motor function) and 66 (intact motor function). The reported minimal clinically important difference (MCID) for the FMA-UE is 5.2 points (47).

Secondary outcome measures assessed upper limb capacity on activity level according to the International Classification of Functioning, Disability and Health (ICF) (48). The ARAT measures motor performance in 4 different subscales: grasp, grip, pinch and gross movement, with a maximum score of 57, reflecting good motor performance. The reported MCID for the ARAT is 6 points (49). The SULCS is a hierarchical 10-item scale evaluating upper limb capacity in functional tasks, with the total score corresponding to the number of tasks the patient is able to execute. The BBT evaluates gross manual dexterity by the number of blocks of 1cm^3^ that could be moved between two boxes within one minute. The reported MCID for the BBT is 6 blocks (49). The JEBSEN assesses manual dexterity in six unimanual tasks, by means of movement time (seconds), with lower scores indicating better capacity. Finally, the RMA-A assesses motor performance in the arm using 15 hierarchically ordered items that are scored dichotomously (0-1). Higher score reflects better motor performance. Adequate psychometric properties are established for all outcome measures (49). Measures of study therapy compliance included the number of sessions completed, both for the BOOST and CONTROL intervention.

### 2.5 Statistical analysis

Clinical and demographic characteristics of participants at baseline were displayed as frequencies with percentage and medians with interquartile range (IQR), whichever appropriate. Clinical and baseline characteristics from patients that completed the study were compared to characteristics of patients that dropped-out during follow-up, by using Mann-Whitney U tests and Chi square tests. To compare characteristics and baseline data, including therapy compliance and usual care, between the immediate and delayed group, Mann-Whitney U tests and Chi square tests were used. Between-group differences in change over time in all outcome measures between BASELINE and POST 1 and between POST 1 and POST 2 were assessed using independent-samples Mann-Whitney U tests. For the outcome measures with reported MCIDs, we explored whether individual change scores exceeded the MCID threshold. P-values were considered statistically significant at the 0.05 level. Correction for multiple testing was not performed due to the pilot design. This is a pilot study and therefore, no sample size calculation was conducted. All statistical analyses were performed using PAWS statistics by SPSS, version 18.

## 3 Results

### 3.1 Participants

A total of 18 participants were randomized and allocated to the immediate group (IG: n=10) or delayed group (DG: n=8). At BASELINE and POST 1, data from all participants were available. At the final measurement (POST 2), 5 patients dropped out from the IG and 1 patient from the DG was lost during follow-up. Patients that dropped out during follow-up were not different from the patients that completed the study for age (p=0.261), gender (p=0.710), time post stroke (p=0.075), lateralization (p=0.502), type of stroke (p=0.482), score on Barthel index (p=0.100), score on Montreal cognitive assessment (p=0.479), and score on star cancellation test (p=0.076). The flow diagram, including reasons for drop-out can be found in figure 1.

Table 1 summarizes the demographic and clinical characteristics at baseline for the participants. Median age (IQR) at stroke onset was 65.3 years (51.8-72.8) and 72% of the participants was female. Participants entered the study at a median (IQR) of 8.6 weeks (5-12.4), 72% experienced ischemic stroke and 44% had left hemiplegic stroke. Baseline clinical assessment showed moderate to severe dependence in activities of daily living and moderate cognitive impairments. Demographic and clinical characteristics of patients from the IG were not significantly different from characteristics of patients from the DG. However, a tendency can be noticed in time post stroke (p=0.083) and cognitive functioning (p=0.099) with patients from the DG entering the study at a relatively later phase post stroke and experiencing relatively more cognitive impairments.

### 3.2 Therapy compliance and usual care

Therapy compliance was moderate to high and comparable between both groups (p=0.343) with completed study therapy hours (including both BOOST and CONTROL) at the end of the study ranging between 40 and 45 hours for the IG and between 38-43 for the DG. Additionally, the amount of usual care was comparable between both groups (p=0.515), with a median (IQR) of 52.8 (45.8-57.5) hours in the IG and 54.5 (50.5-58.4) hours in the DG over the first 4 weeks of the study. Put differently, patients received between 1.8 and 3.5 hours of usual care per day, besides the additional study therapy program. No adverse events were experienced during the study.

### 3.3 Clinical outcome

Table 2 shows the time course of the different outcome measures for the IG and DG. Median baseline scores of the 2 groups did not differ significantly for any of the outcome measures (p=0.052 to p=1). The primary outcome measure was the change in upper limb impairment measured by FMA-UE, from BASELINE to POST 1 (after 4 weeks of training). A significant between-group difference (p=0.003) was found with median (IQR) change scores of 9 (7.3-16.3) and 0.5 (−2.8-3) for the IG and DG, respectively. Conversely, between POST 1 and POST 2, patients in the DG improved with a median (IQR) of 8 (0-11) points compared to 1 (−3-2.5) point in the IG, although this difference did not reach significance (p=0.103).

On upper limb capacity level, there is a significant difference (p=0.033) between both groups for the change in SULCS scores between BASELINE and POST 1, and a trend (p=0.075) for the change in ARAT scores. In these first 4 weeks of additional therapy, patients in the IG improved with a median (IQR) of 2 (1-3) points on the SULCS and 12.5 (1.3-17.8) points on the ARAT, whereas the DG improved with median scores (IQR) of 1 (0.3-1) for the SULCS and 1.5 (−1-8.5) for the ARAT. Between POST 1 and POST 2, patients in the DG tend to show larger improvements in ARAT and SULCS when compared to the IG, however, between-group comparisons did not reach significance (p=0.240 – p=0.416). Finally, no significant differences between groups were found for BBT, JEBSEN and RMA-A (p=0.120 – p=0.673).

For the outcome measures with reported MCIDs, we explored whether individual change scores exceeded the MCID threshold (Figure 2-4). For the FMA-UE, 80% of patients in the IG exceeded the threshold of 5.2 points between BASELINE and POST 1, whereas none of the patients in the DG did (Figure 2a). Between POST 1 and POST 2, patients of the IG remained stable as none of them exceeded the threshold again. Four out of seven patients from the DG exceeded the threshold in this last four weeks of the study (Figure 2b). A similar pattern is seen for the ARAT (Figure 3). Between BASELINE and POST 1, 70% of patients from the IG and 25% of patients from the DG showed improvements beyond the MCID, and 40% vs. 57% between POST 1 and POST 2, respectively. Finally, for the BBT, 60% in the IG and 37.5% in the DG exceeded the MCID of 6 blocks in the first four weeks (Figure 4a). Between POST 1 and POST 2, patients of the IG remained stable as none of them exceeded the threshold again. three out of seven patients from the DG exceeded the threshold in this last four weeks of the study (Figure 4b).

**Figure 2.**
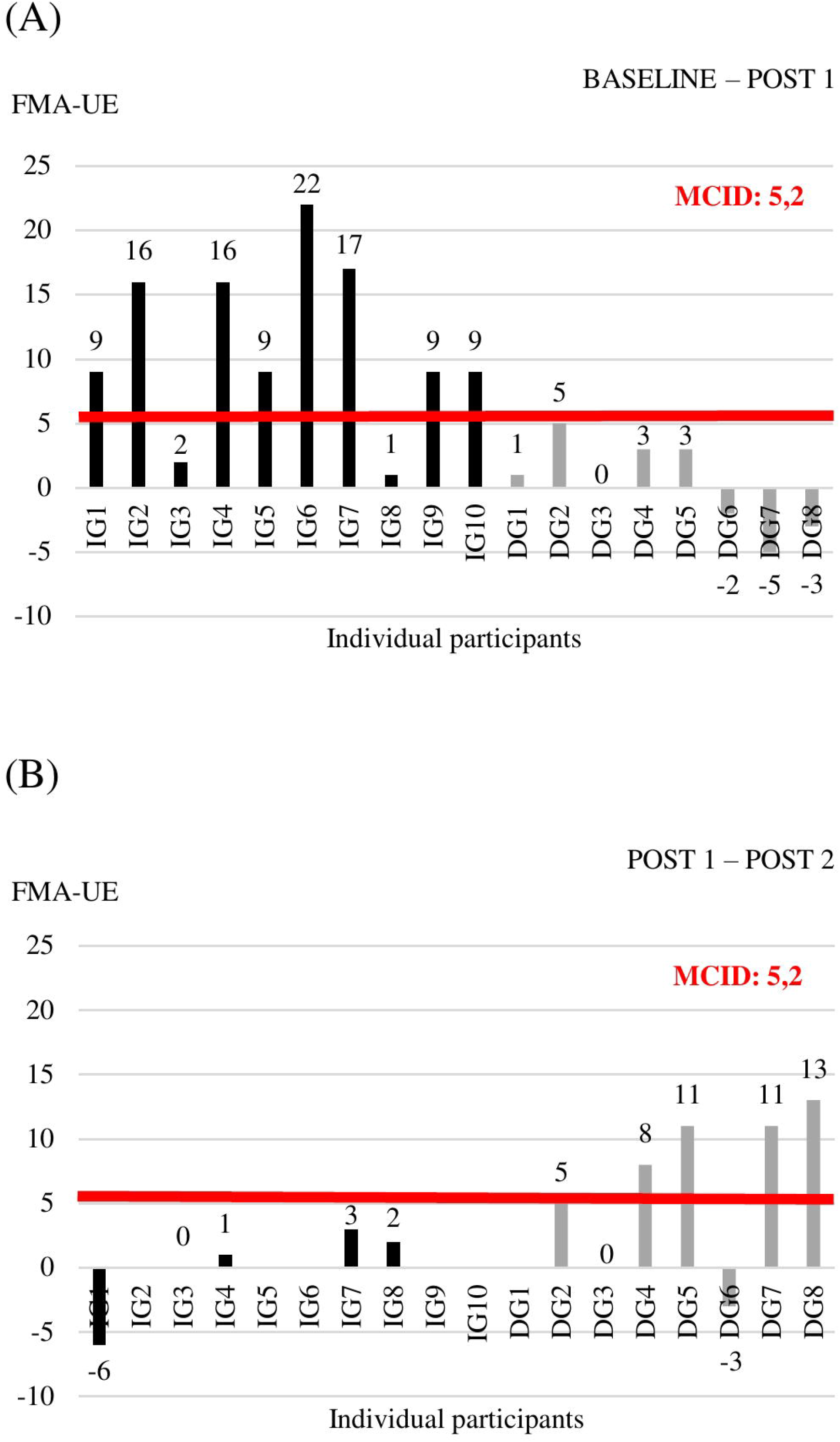
Individual change scores in FMA-UE between BASELINE and POST 1 (A) and between POST 1 and POST 2 (B)

**Figure 3.**
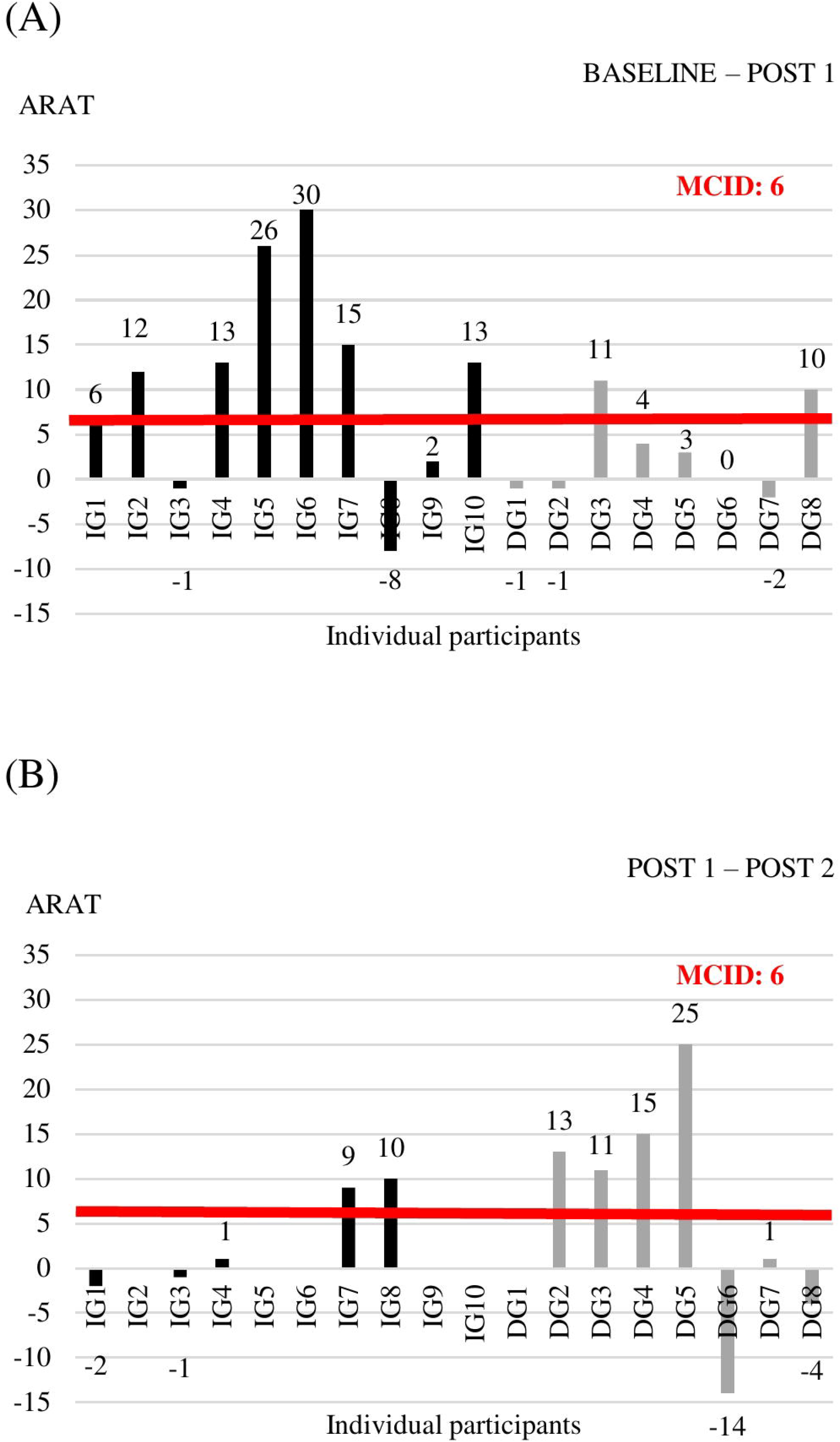
Individual change scores in ARAT between BASELINE and POST 1 (A) and between POST 1 and POST 2 (B)

**Figure 4.**
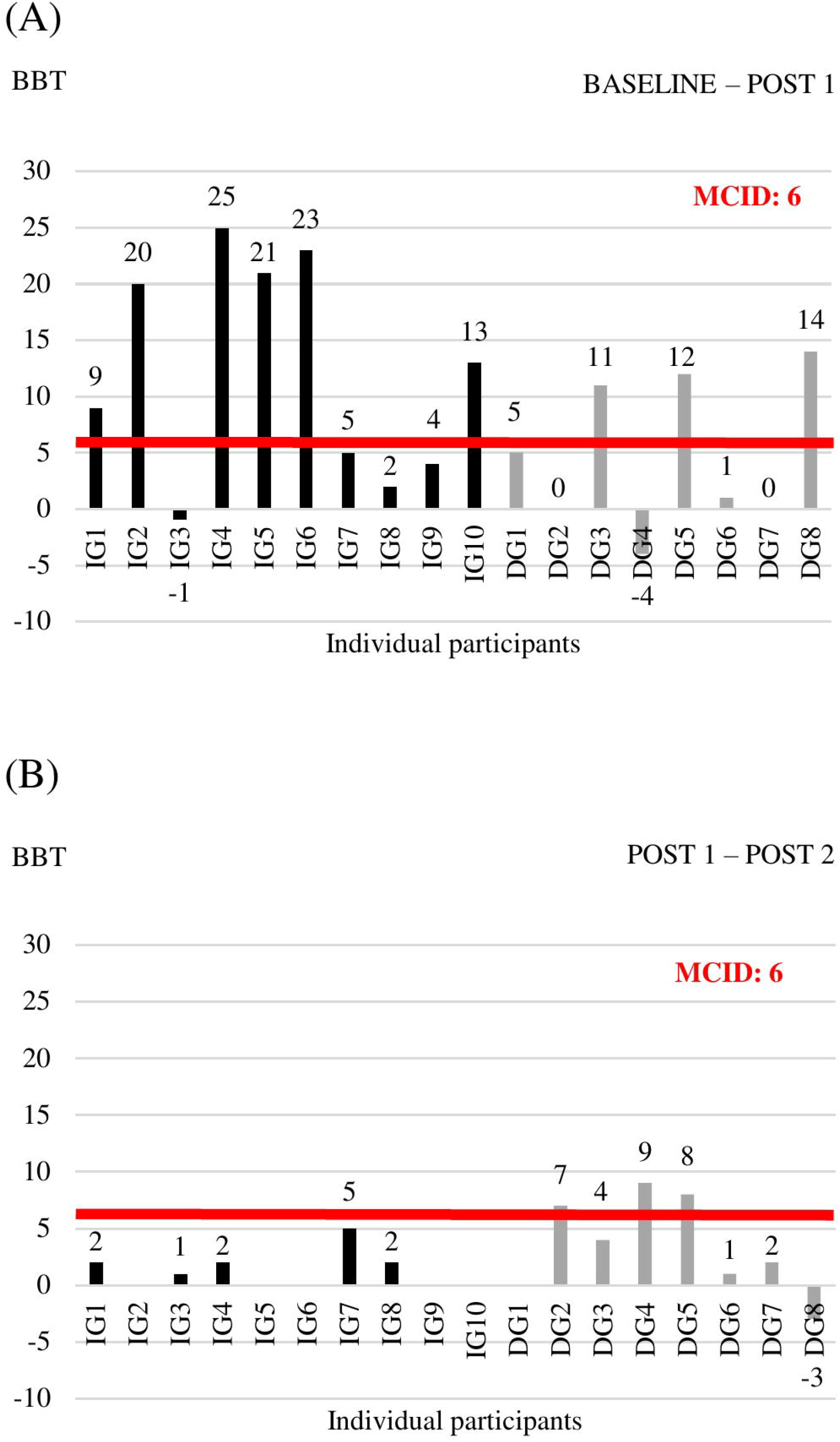
Individual change scores in BBT between BASELINE and POST 1 (A) and between POST 1 and POST 2 (B)

## 4 Discussion

This pilot randomized controlled study revealed that our intensive arm-hand BOOST program is feasible and safe when embedded in the inpatient rehabilitation phase post stroke. Also, this study suggests that the BOOST program is beneficial for improving upper limb motor function and activities. Finally, it may be that early delivery of the BOOST program is superior when compared to late delivery post stroke, with more patients exceeding the minimal clinical important difference of upper limb outcomes when the BOOST program is provided in the early sub-acute phase.

Results of the study regarding feasibility and safety of providing extra hours of focused therapy showed moderate to high compliance to the study therapy and no adverse events. Additionally, in this study a range of 1.8 to 3.5 hours of usual care therapy per day was provided, consisting of physiotherapy and occupational therapy, supplemented by speech- and language therapy, psychological therapy and sports therapy. Therefore, patients practiced up to 5 hours per day during the course of the study. These results are in line with previous studies showing that high amounts of therapy and high intensities are well tolerated (31-33,50), despite the skepticism both from a patients’ and organizational perspective. With regard to tolerability of high intensity training, Waddell et al. (50) showed that patients with subacute stroke can achieve on average 289 repetitions per hour of task-specific UE training and higher doses were associated with better outcomes on the ARAT (50).

This study showed the combined, positive effect of a high-dose, focused upper limb program including principles of neurophysiological recovery in the sub-acute phase post stroke, on upper limb impairment and functional activities. Large improvements were found on both impairment and activity level, with a median improvement of 9 points of the FMA-UE and 12.5 points on the ARAT after 4 weeks of early BOOST therapy. Our results are in line with the magnitude of changes reported in recent studies with patients in the chronic phase post stroke, albeit with smaller improvements in those studies. In the cohort study of Ward et al. (31), the Queen Square Upper Limb Neurorehabilitation program was provided to 224 patients in the chronic phase post stroke. A median increase of 6 points was found for the FMA-UE and 6 points for the ARAT after 90 hours of high intensity arm and hand treatment, provided over 3 weeks. Continued improvements were shown up to six months after treatment (31). Similarly, a mean 10-point increase in FMA-UE scores was found in the study of Daly et al. (32), which is a replication of a previously published study of McCabe et al. (33), who provided 300 hours of upper limb motor learning combined with the use of technology to chronic stroke patients in a 12-week program. Gains in motor function were maintained up to 3 months follow-up (32).

Very recently, Krakauer et al. (30) reported significant improvements in ARAT scores (mean difference 7.33 ± 2.88), and not in Fugl-Meyer scores (mean difference 1.44 ± 2.57,), when intensifying the rehabilitation training program in the sub-acute phase post stroke by using a novel exploratory neuro-animation therapy, when compared to the usual care (30). These slightly contradictory findings when compared to our results, might be explained by the content of the therapy program. In the study of Krakauer et al. (30) the exploratory neuro-animation therapy is performed using the Armeo^®^Power, an exoskeleton device which allows 3D-movements of shoulder, elbow and wrist. Therefore, the program has less focus on improving distal hand impairment as well as adequate postural control and core stability, correct scapula setting and efficient scapular humeral rhythm as prerequisites for recovery of voluntary selective movements.

The positive effect of our BOOST program was very clear for most of the patients, with a compelling majority of participants improving with clinical important steps in measures of impairment and activities. However, there are still a few non-responders to the BOOST program. When looking at individual data (Figure 2a, Figure 3a), one might notice 2 participants in the IG that did not improve beyond the MCID thresholds after four weeks of BOOST therapy. One participant (IG3) showed communication problems and low motivation during therapy. It is known from previous literature that stroke-related psychological issues negatively influence rehabilitation and outcomes through a reduction in compliance to the exercise program, low dedication and concentration during the sessions, increased fatigue levels, and potentially less motivation (51). The other non-responder (IG8) developed a limiting tremor in the hand, which impacted slightly on the FMA-UE assessment but had a large impact on the ARAT and BBT assessments.

Interestingly, our study also suggests that timing of the delivery of the program matters. Although all patients in the study received exactly the same amount of therapy and the same content, the order in which the two therapies (BOOST and CONTROL) were applied differed between the IG and the DG. Results showed that early delivery of the BOOST program was better compared to late delivery, with 80% of patients in the IG improving beyond the MCID of the FMA-UE, compared to 57% in the DG. This can be related to the critical window of opportunity for recovery when the brain is most responsive to sensorimotor experience after injury, typically seen in the first 3 months post stroke (9,52,53). However, it is also important to notice that patients from the DG entered the study on average at a later phase post stroke with a median of 11.6 weeks post stroke, compared to 7.6 weeks post stroke in the IG, although the difference was non-significant. This might have contributed to the difference between both groups.

Up to now, studies concentrated mainly on offering higher doses of upper limb therapy in the chronic phase post stroke, whereas our study adds to the body of knowledge regarding the combined effect of a more focused upper limb program, including principles of neurophysiological recovery, provided at high intensities, embedded within the early inpatient rehabilitation phase post stroke. Additionally, it was the first study directly comparing the effect of early versus late delivery of the program in the subacute phase post stroke. However, some limitations of the study need to be recognized. First, in this pilot study participants entered the study between 15 and 367 days stroke, which covers a broad time window for inclusion of patients. Of our sample of 18 patients, 72% was in the early sub-acute phase (up to three months post stroke) and 17% was in the late sub-acute phase (between three and six months). Additionally, although the difference was non-significant, patients from the DG entered the study on average at a later phase post stroke compared to IG. Since this pilot study has only a restricted sample size, it is recommended to stratify patients according to timing post stroke when setting up a future randomized controlled trial. Secondly, due to the restricted sample size, we were unable to unravel key characteristics of non-responders to the BOOST therapy. Third, there was a large range in amount of usual care (between 1.8 and 3.5 hours of usual care per day), provided to the participants. Moreover, there is no further information concerning the content of the different usual care programs, which would have been useful in exploring the total amount of therapy time for the upper limb, provided by different therapists. Finally, there was no follow-up measurement after the end of the treatment. It would be of interest whether the difference between the IG and DG would be retained up to several months after the intervention.

Future research is needed and justified, including a phase III trial of patients receiving the BOOST therapy in the sub-acute phase post stroke, stratified according to timing post stroke. Also, a follow-up period of several months after treatment is needed in order to further examine the long-term effects. Additionally, patient experience should be monitored. Finally, more detailed insights in the cost-effectiveness of the BOOST program is warranted. Benefits might be first seen in improvements in activities of daily living. These gains might then result in less caregiver need, reducing cost of care. Additionally, benefits might also be reflected in increased quality of life and reduced anxiety and depression.

In summary, our findings highlight that an intensive specific arm-hand BOOST program, on top of usual care, is feasible and safe in the subacute phase post stroke and suggests a positive, clinical meaningful effect on upper limb function and activity, especially when delivered in the early sub-acute phase post stroke.

## Data Availability

The data associated with the paper are not publicly available but are available from the corresponding author on reasonable request.

## 6 Tables

**Table 1.**
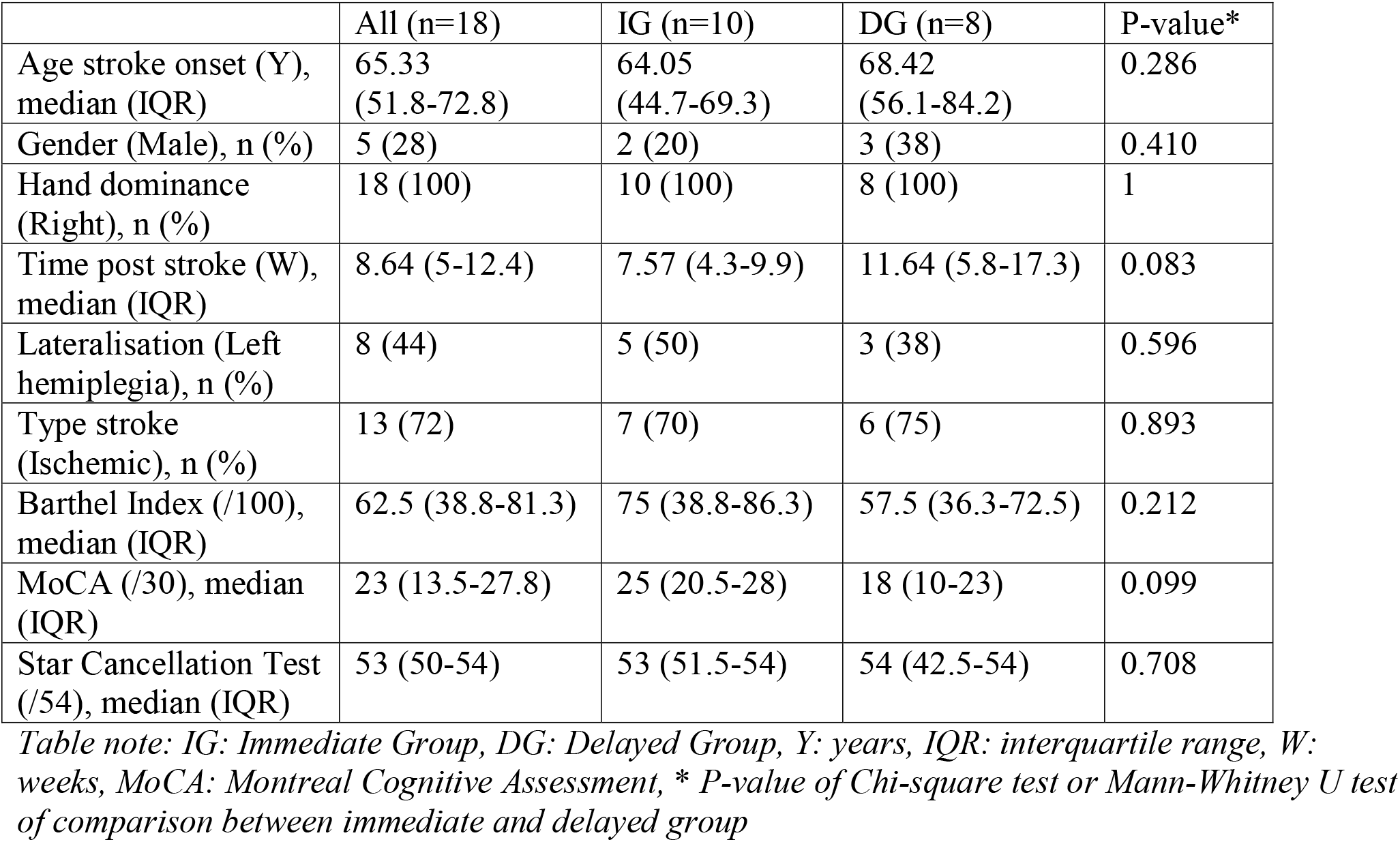
Patients’ demographic and clinical information at baseline

**Table 2.** Time course of outcome measures for the immediate group and delayed group

|  |  | FMA-UE | ARAT | SULCS | BBT | JEBSEN | RMA-A |
| --- | --- | --- | --- | --- | --- | --- | --- |
| BASELINE:<br>median<br>(IQR) | IG<br>(n=10) | 37<br>(26.25-44) | 25.5<br>(15.75-33.75) | 5 (2.75-6) | 10.5<br>(0.75-19.25) | 525 (365-788) | 4.5 (1.75-6.75) |
|  | DG<br>(n=8) | 30<br>(22.75-45.25) | 19<br>(14.25-33.75) | 3 (2-4.5) | 10 (2.25-13) | 663 (407-767) | 1.5 (1-3.5) |
|  | P-value* | 0.477 | 0.593 | 0.259 | 1 | 0.790 | 0.052 |
| POST 1:<br>median<br>(IQR) | IG<br>(n=10) | 51.5<br>(41.5-54.5) | 39 (26.5-46) | 7 (4-8) | 22.5 (11-29.25) | 265 (183-367) | 8 (3-11.25) |
|  | DG<br>(n=8) | 29.5<br>(25.75-41.75) | 19 (15-35.25) | 4 (2.25-5.75) | 12.5<br>(6.75-19.25) | 486 (412-510) | 3 (1.25-4) |
|  | P-value* | **0.016** | 0.075 | 0.058 | 0.168 | **0.026** | **0.044** |
| POST 2:<br>median<br>(IQR) | IG<br>(n=5) | 53 (31.5-56) | 50 (26-53.5) | 7 (5.5-9) | 23 (12.5-38) | 244 (141-366) | 8 (2.5-12.5) |
|  | DG<br>(n=7) | 36 (27-48) | 33 (20-41) | 4 (3-6) | 16 (13-28) | 392 (232-480) | 3 (1-8) |
|  | P-value* | 0.370 | 0.193 | 0.118 | 0.626 | 0.223 | 0.192 |
| BASELINE<br>– POST 1:<br>median<br>(IQR) | IG<br>(n=10) | 9 (7.25-16.25) | 12.5<br>(1.25-17.75) | 2 (1-3) | 11 (3.5-21.5) | -153 (-<br>494--86) | 1.5 (0-5.5) |
|  | DG<br>(n=8) | 0.5 (-<br>2.75-3) | 1.5 (-1-8.5) | 1 (0.25-1) | 3 (0-11.75) | -170 (-<br>241--17) | 1 (0-2) |
|  | P-value* | **0.003** | 0.075 | **0.033** | 0.120 | 0.424 | 0.490 |
| POST 1 –<br>POST 2:<br>median<br>(IQR) | IG<br>(n=5) | 1 (-3-2.5) | 1 (-1.5-9.5) | 0 (0-1) | 2 (1.5-3.5) | -11 (-<br>81.5-1) | 0 (-1-1.5) |
|  | DG<br>(n=7) | 8 (0-11) | 11 (-4-15) | 1 (0-1) | 4 (1-8) | -74 (-<br>118--35) | 1 (0-1) |
|  | P-value* | 0.103 | 0.416 | 0.240 | 0.456 | 0.167 | 0.673 |

Table note: IG: Immediate Group, DG: Delayed Group, FMA-UE: Fugl-Meyer assessment upper extremity, ARAT: Action research arm test, SULCS: Stroke upper limb capacity scale, BBT: Box & block test, JEBSEN: Jebsen Taylor hand function test, RMA-A: Rivermead motor assessment – arm subscale, \* P-value of Mann-Whitney U test of comparison between immediate and delayed group

## 10 Authors’ contribution

SM, GV and MM have given substantial contributions to the conception and the design of the manuscript. All authors have given substantial contribution to acquisition, analysis and interpretation of the data. SM drafted the manuscript, and all authors revised it critically. All authors read and approved the final version of the manuscript.

## 12 Acknowledgements

The authors would like to thank all therapists from the inpatient rehabilitation unit of Jessa Hospital, Rehabilitation Campus Sint-Ursula in Belgium who provided all study therapy sessions.

