## Supplementary tables for "Arm-hand boost therapy during inpatient stroke rehabilitation: a pilot randomized controlled trial"

### *Supplementary Material*

**Supplementary Table 1**

**CONSORT 2010 checklist of information to include when reporting a randomised trial**

| Section/Topic | Item No | Checklist item | Reported on page No |
|---|---|---|---|
| **Title and abstract** | | | |
| | 1a | Identification as a randomised trial in the title | 1 |
| | 1b | Structured summary of trial design, methods, results, and conclusions (for specific guidance see CONSORT for abstracts) | 2 |
| **Introduction** | | | |
| Background and objectives | 2a | Scientific background and explanation of rationale | 3-4 |
| | 2b | Specific objectives or hypotheses | 3-4 |
| **Methods** | | | |
| Trial design | 3a | Description of trial design (such as parallel, factorial) including allocation ratio | 5 |
| | 3b | Important changes to methods after trial commencement (such as eligibility criteria), with reasons | N.A. |
| Participants | 4a | Eligibility criteria for participants | 5 |
| | 4b | Settings and locations where the data were collected | 5 |
| Interventions | 5 | The interventions for each group with sufficient details to allow replication, including how and when they were actually administered | 5-6 |
| Outcomes | 6a | Completely defined pre-specified primary and secondary outcome measures, including how and when they were assessed | 6-7 |
| | 6b | Any changes to trial outcomes after the trial commenced, with reasons | N.A. |
| Sample size | 7a | How sample size was determined | N.A. |
| | 7b | When applicable, explanation of any interim analyses and stopping guidelines | N.A. |

| | | | |
|---|---|---|---|
| Randomisation: | | | |
| Sequence generation | 8a | Method used to generate the random allocation sequence | 5 |
| | 8b | Type of randomisation; details of any restriction (such as blocking and block size) | 5 |
| Allocation concealment mechanism | 9 | Mechanism used to implement the random allocation sequence (such as sequentially numbered containers), describing any steps taken to conceal the sequence until interventions were assigned | 5 |
| Implementation | 10 | Who generated the random allocation sequence, who enrolled participants, and who assigned participants to interventions | 5 |
| Blinding | 11a | If done, who was blinded after assignment to interventions (for example, participants, care providers, those assessing outcomes) and how | 7 |
| | 11b | If relevant, description of the similarity of interventions | N.A. |
| Statistical methods | 12a | Statistical methods used to compare groups for primary and secondary outcomes | 7 |
| | 12b | Methods for additional analyses, such as subgroup analyses and adjusted analyses | 8 |
| **Results** | | | |
| Participant flow (a diagram is strongly recommended) | 13a | For each group, the numbers of participants who were randomly assigned, received intended treatment, and were analysed for the primary outcome | 9 + figure 1 |
| | 13b | For each group, losses and exclusions after randomisation, together with reasons | Figure 1 |
| Recruitment | 14a | Dates defining the periods of recruitment and follow-up | 5 |
| | 14b | Why the trial ended or was stopped | N.A. |
| Baseline data | 15 | A table showing baseline demographic and clinical characteristics for each group | Table 1 |
| Numbers analysed | 16 | For each group, number of participants (denominator) included in each analysis and whether the analysis was by original assigned groups | 9, Table 1+2 |
| Outcomes and estimation | 17a | For each primary and secondary outcome, results for each group, and the estimated effect size and its precision (such as 95% confidence interval) | 9,10,Table 2 |
| | 17b | For binary outcomes, presentation of both absolute and relative effect sizes is recommended | 10 |
| Ancillary analyses | 18 | Results of any other analyses performed, including subgroup analyses and adjusted analyses, distinguishing pre-specified from exploratory | N.A. |
| Harms | 19 | All important harms or unintended effects in each group (for specific guidance see CONSORT for harms) | 9 |
| **Discussion** | | | |

| | | | |
|---|---|---|---|
| Limitations | 20 | Trial limitations, addressing sources of potential bias, imprecision, and, if relevant, multiplicity of analyses | 12 |
| Generalisability | 21 | Generalisability (external validity, applicability) of the trial findings | 12 |
| Interpretation | 22 | Interpretation consistent with results, balancing benefits and harms, and considering other relevant evidence | 11-13 |
| **Other information** | | | |
| Registration | 23 | Registration number and name of trial registry | 5 |
| Protocol | 24 | Where the full trial protocol can be accessed, if available | N.A. |
| Funding | 25 | Sources of funding and other support (such as supply of drugs), role of funders | 19 |

**Supplementary Table 2**

TIDieR Template for Intervention Description and Replication

**The TIDieR (Template for Intervention Description and Replication) Checklist:**
Information to include when describing an intervention and the location of the information

| Item number | Item | Where located | |
|---|---|---|---|
| | | Primary paper (page number) | Other † (details) |
| | **BRIEF NAME** | | |
| **1.** | **Provide the name or a phrase that describes the intervention.**<br><br>Intensive BOOST program for the upper limb | 6 | ______________ |
| | **WHY** | | |
| **2.** | **Describe any rationale, theory, or goal of the elements essential to the intervention.**<br><br>The focussed therapy program for the upper limb is built upon knowledge of neurophysiological recovery post stroke to ensure that specific therapy goals are set for the correct patient at each stage in recovery. The prerequisites for recovery of voluntary selective movements of the upper limb in patients post stroke include a large proximal component such as adequate postural control and core stability, correct scapula setting, efficient scapular humeral rhythm and selective recruitment of reach-related musculature. Besides the proximal component, specific attention is needed towards the rehabilitation of the hand, including restoring the different arches of the hand to provide a stable base and correct alignment as a prerequisite for dexterity and hand shape modulation while reaching to objects. Additionally, considerable agreement exists about the importance of high dose (time in rehabilitation, or amount of repetitions) and intensity (dose per session) of upper limb therapy. Therefore, the combined effect of of a more focussed program, including principles of neurophysiological recovery, provided at high intensities in the sub-acute phase post stroke is researched. | 3 | _____________ |

| | | | |
|---|---|---|---|
| | **WHAT** | | |
| **3.** | **Materials: Describe any physical or informational materials used in the intervention, including those provided to participants or used in intervention delivery or in training of intervention providers. Provide information on where the materials can be accessed (e.g. online appendix, URL).**<br><br>The material used in a BOOST session includes standard rehabilitation material such as different sizes of balls, cones, hoops, beads, and daily objects. | 6 | ____________ |
| **4.** | **Procedures: Describe each of the procedures, activities, and/or processes used in the intervention, including any enabling or support activities.**<br><br>The specific intensive boost program for the upper limb (BOOST) is focused around 5 topics: scapula-setting, core-stability training in relation to reaching, training of external shoulder rotation and elbow extension (movements with 30-60° flexion/abduction in shoulder), fine manipulation or dexterity training and integration in complex ADL tasks. For each of those topics, a list of example exercises was created that could be used depending on the individual abilities of the patient, including a gradual increase in levels of difficulty. Each of the interventions is tailored to the individual patient, based upon the ongoing assessment using the Model of Bobath Clinical Practice, discussion within the group of therapists and individual treatment goals of the patient.<br><br>A typical BOOST session would include exercises and tasks with the aim of getting a proper alignment of the upper limb (eg. eccentric mobilisation of shortened muscles, realignment of scapulohumeral joint), updating orientation (eg. light touch, proprioceptive accuracy), activating the upper limb (eg. scapular stability, weight bearing, activating the intrinsic muscles of the hand, strengthening, reaching, grasping, pinching) and training bilateral limb coordination during functional activities (eg. cooking/eating, dressing, grooming, cleaning, gardening).<br><br>The group sessions were executed with a maximum of four participants, under supervision of two experienced therapists (physiotherapist and/or occupational therapist). Additionally, patients exercise one hour per week using the Armeo®Power (Hocoma AG, Switzerland), an upper limb exoskeleton device. This allowed practice of 3D arm | 6 | ____________ |

movements with six actuated degrees-of-freedom. Participants performed exercises at high intensities in the active-assisted mode by using motivational games. A licensed occupational therapist was present throughout each session and provided feedback to encourage normal (non-synergistic) movement patterns at all times.

**WHO PROVIDED**

**5.** **For each category of intervention provider (e.g. psychologist, nursing assistant), describe their expertise, background and any specific training given.** 6 ______________

The group sessions were executed under supervision of two experienced therapists (physiotherapist and/or occupational therapist). Individual exercises performed with the Armeo®Power (Hocoma AG, Switzerland), were supervised by a licensed occupational therapist.

**HOW**

**6.** **Describe the modes of delivery (e.g. face-to-face or by some other mechanism, such as internet or telephone) of the intervention and whether it was provided individually or in a group.** 6 ______________

The BOOST intervention consist of 20 one-hour group sessions (with a maximum of four patients in each group), face-to-face delivered by two experienced therapists (physiotherapist and/or occupational therapist) and one hour of individual face-to-face therapy per week, supervised by an occupational therapist.

**WHERE**

**7.** **Describe the type(s) of location(s) where the intervention occurred, including any necessary infrastructure or relevant features.** 5 ______________

The intervention was provided in the inpatient rehabilitation unit of Jessa Hospital, Rehabilitation Campus Sint-Ursula in Belgium.

| | | | |
|---|---|---|---|
| | **WHEN and HOW MUCH** | | |
| **8.** | **Describe the number of times the intervention was delivered and over what period of time including the number of sessions, their schedule, and their duration, intensity or dose.**<br><br>The BOOST intervention consist of 20 one-hour group sessions (spread over 4 weeks, 5 days a week) and one hour of individual therapy per week. In total, the study intervention includes 24 hours of therapy, provided over a period of 4 weeks. | 5 | ____________ |
| | **TAILORING** | | |
| **9.** | **If the intervention was planned to be personalised, titrated or adapted, then describe what, why, when, and how.**<br><br>For each of the five topics of the BOOST program, a list of example exercises was created that could be used depending on the individual abilities of the patient, including a gradual increase in levels of difficulty. Each of the interventions is tailored to the individual patient, based upon the ongoing assessment using the Model of Bobath Clinical Practice, discussion within the group of therapists and individual treatment goals of the patient. | 6 | ____________ |
| | **MODIFICATIONS** | | |
| **10.**[ǂ] | **If the intervention was modified during the course of the study, describe the changes (what, why, when, and how).**<br><br>No modifications during the course of the study. | N.A. | ____________ |
| | **HOW WELL** | | |
| **11.** | **Planned: If intervention adherence or fidelity was assessed, describe how and by whom, and if any strategies were used to maintain or improve fidelity, describe them.**<br><br>Treating therapists recorded the presence of the patient in each of the therapy sessions. Study therapy sessions were incorporated into the usual care treatment schemes of the patients to improve fidelity. | 5 | ____________ |

| | | | |
|---|---|---|---|
| **12.**‡ | **Actual: If intervention adherence or fidelity was assessed, describe the extent to which the intervention was delivered as planned.**<br><br>Therapy compliance was moderate to high and comparable between both groups with completed study therapy hours (including both BOOST and CONTROL) at the end of the study ranging between 40 and 45 hours for the Immediate Group and between 38 and 43 for the Delayed Group (out of 48 hours in total). | 9 | ____________ |
